# A sensitive multiplex RT-qPCR assay to detect SARS-CoV-2 in respiratory samples

**DOI:** 10.1101/2025.03.31.25323652

**Authors:** Alice Halliday, Begonia Morales-Aza, Kaltun Duale, Philippa Lait, Bristol Vaccine Centre Microbiology & Molecular Laboratories, Rachel Milligan, Stephanie Hutchings, Hannah Pymont, Barry Vipond, Peter Muir, Christy Waterfall, Jane Coghill, Jeremy Carr, Emma Plested, Jennifer Oliver, Katy Fidler, Caroline Relton, Matthew D Snape, the ‘Be on the Team’ Investigators, Andrew D. Davidson, Christopher Helps, Adam Finn

**Affiliations:** Bristol Vaccine Centre, University of Bristol; School of Cellular and Molecular Medicine, University of Bristol.; Langford Vets, University of Bristol; School of Veterinary Sciences, University of Bristol; UK Health Security Agency (UKSHA); Genomics Facility, Faculty of Health & Life Sciences, University of Bristol; Oxford Vaccine Group, University of Oxford Department of Paediatrics; Population Health Sciences, Bristol Medical School, University of Bristol, UK; Brighton and Sussex Medical School, Falmer, UK; Royal Alexandra Children’s Hospital, University Hospital Sussex NHS Foundation Trust; MRC Integrative Epidemiology Unit, Population Health Sciences, Bristol Medical School, University of Bristol, UK; 12-London School of Hygiene & Tropical Medicine, London, UK; Oxford NIHR Biomedical Research Centre; Paediatric Immunology and Infectious Diseases, Bristol Royal Hospital for Children

## Abstract

We developed an in-house RT-qPCR assay for the detection of SARS-CoV-2 from different sample types. Novel primer and probe sets were designed to amplify 2 regions of the nucleocapsid (N) gene. Two dual-target multiplex assays were then evaluated for performance in detection of SARS-CoV-2 in known samples. An assay combining one novel primer pair for N (N6), a published region for the envelope protein (E) and an extraction control target (MS2), was found to have good efficiency and a low limit of detection, and to be sensitive and specific for SARS-CoV-2 in a sample set of known positive and negative clinical samples. Assay performance was maintained for the main variants of concern (Alpha, Delta, Gamma and Omicron BA.1, BA.2, BA4, JN.1). The assay was used to test for SARS-CoV-2 in throat swab samples collected in schools from teenagers in the UK as part of an observational study of two Meningococcal B Vaccines in Jan-March 2020. The low prevalence of positives identified (0.44-0.47% in two weeks of March 2020) suggests that there was minimal circulation of SARS-CoV-2 in UK schools prior to the first lockdown. The N6/E/MS2 assay has since been used to detect SARS-CoV-2 in samples from multiple studies. Continual monitoring will ensure that performance of this novel multiplex assay is maintained as novel sub-variants emerge.

## Introduction

The emergence of severe acute respiratory syndrome coronavirus 2 (SARS-CoV-2) which causes coronavirus infectious disease 2019 (COVID-19) has led to a pandemic with severe health, social and economic consequences globally. Some early attempts to establish diagnostic PCR assays to detect SARS-CoV-2 using a range of gene targets including those encoding the RNA-dependent RNA polymerase (RdRp) and nucleocapsid (N), envelope (E) and Spike (S) proteins (1, 2) were only moderately successful with relatively low sensitivity and significant numbers of false negatives. Subsequently a range of high performing assays have been developed and deployed by government and academic labs and numerous commercial assays have also become available.

Lateral flow antigen tests, which are less sensitive but reasonably predictive of infectiousness have the important advantages of low cost, rapid results with no requirement for laboratory processing and have been widely deployed to support public health efforts to control spread of the infection. Nevertheless, RT-qPCR remains the gold standard testing methodology for individual diagnosis of SARS-CoV-2 infection with high sensitivity and specificity.

Here we describe development, evaluation and deployment of a novel SARS-CoV-2 RT-qPCR assay which uses Envelope (E) and Nucleocapsid (N) gene targets and delivers high technical and diagnostic performance. We also present data from the testing of throat swabs samples collected from teenagers in England in early 2020, at the very start of the UK SARS-CoV-2 epidemic.

## Methods

### Samples, Ethics and Consent

Frozen SARS-CoV-2 positive and negative nasopharyngeal/oropharyngeal swabs (NPS/OPS) in viral transport medium (VTM) collected in early 2020 were taken from individuals undergoing testing for SARS-CoV-2 across the Bristol area. These were tested at Severn Pathology Laboratories, Bristol, UK Health Security Agency (UKHSA), and anonymised remnant samples and laboratory test data were subsequently transferred to the University of Bristol to assist in the validation of two candidate RT-qPCR assays.

For field testing the assay on unknown samples, oropharyngeal swabs (OPS) from healthy 16–17-year-olds enrolled in the “Be on the Team” (BOTT) study were used. This was a large vaccine study in progress at the time evaluating the impact of two protein antigen meningococcal vaccines (Bexsero (GSK) and Trumenba (Pfizer)) on respiratory carriage (3, 4). Signed informed consent was obtained in all cases for surplus material from swab samples to be used for additional research and permission to use them for detection of SARS-CoV-2 was obtained from the ethics committee who first approved the parent study before this work was done (18/SC/0055). Oropharyngeal swabs were collected into 1.5 mL skim milk, tryptone, glucose, and glycerin (STGG) broth (E&O Laboratories) from BOTT at sites in 16 regions across the UK between January and March 2020. These swabs were shipped to the Bristol laboratories for total nucleic acid extraction and RT-qPCR.

Positive control material for SARS-CoV-2 Delta and Omicron strains were obtained with appropriate informed consent via Bristol Biobank (20/WA/0273), from infected adult volunteer donors working in the Biomedical Sciences Building, University of Bristol. Collected specimens were held at 4°C and transported to the laboratories before freezing at −70 °C within 4 hours.

### Extraction

Samples were kept frozen at –70 °C and were defrosted on ice prior to nucleic acid extraction. All samples were first vortexed for 30 seconds, then chemically inactivated by adding 500 µL of L6 Buffer (Severn Biotech, 20-8600-15) to 90 μL sample (on ice), then vortexed for a further 3 seconds and incubated at room temperature for 15 minutes (as per UKHSA Guidelines). Positive and negative samples provided by UKHSA and BOTT samples underwent nucleic acid extraction using the QIAsymphony DSP Virus/Pathogen Mini Kit (QIAGEN), in combination with the QIAsymphony SP instrument, according to the manufacturer’s instructions (Extraction Protocol: Cell_Free 200 with elute volume of 60 µl). Additional extraction protocols (i.e. Macherey Nagel Nucleospin RNA Kit; Kingfisher) were also used and compared for extraction of RNA from control materials and found to provide comparable results to the QIASymphony (results not shown). For all extractions, samples were spiked with the RNA bacteriophage MS2 (genome accession V00642.1, provided by Langford Vets) at a final dilution of 10^-8^ of a lab grown stock to give a CT value of ∼24. This was used as an internal control for the extraction process, RT and qPCR.

### RT-qPCR Assay Design and Initial Optimisation

With the exception of the E gene, where primer and probe sequences were unchanged from those described previously (5), RT-qPCR assays and novel primers were designed for different genes of SARS-CoV-2 as previously described (6), with details provided in Table 1. Novel SARS-CoV-2 primers and probes from different regions of the nucleocapsid (N gene; denoted N2 and N6 (in-house numbering)) were designed using Primer3 (7) using a consensus multiple sequence alignment of 658 SARS-CoV-2 N gene sequences downloaded from GenBank (sequences downloaded April 2020 and Alignments performed in May 2020). Amplicon secondary structure was predicted using Mfold (8).

**Table 1.**
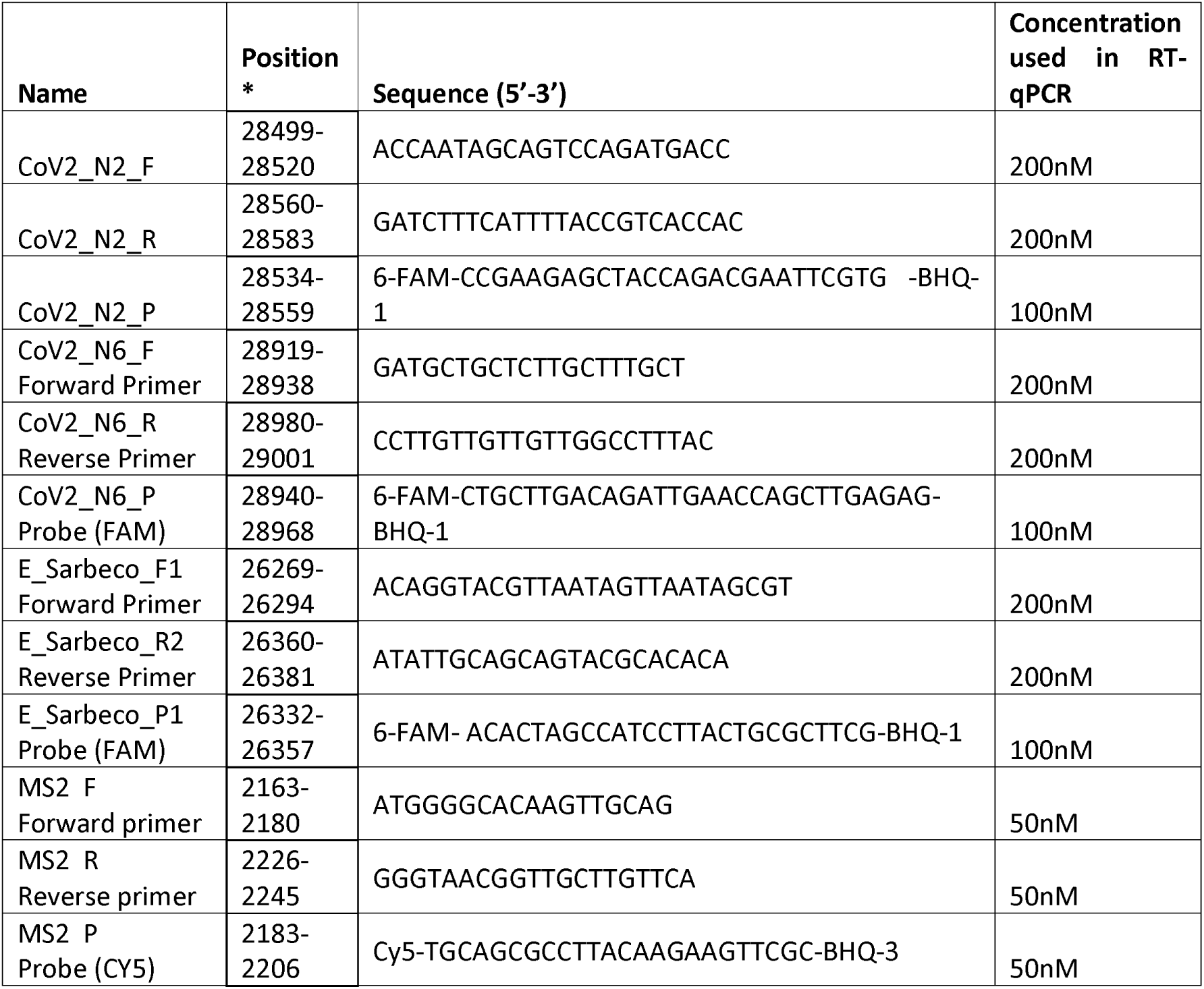
Primer and Probe Sequences. Full sequences and modifications for primers and probes in the SARS-CoV-2 N6/E/MS2 Multiplex Assay as well as the final concentration used in RT-qPCR. * Position indicated is relative to SARS-CoV-2 isolate Wuhan-Hu-1 genome (accession MN908947.3) or MS2 phage genome (accession V00642.1). Probes were labelled with 6-Carboxyfluorescein (6-FAM) and Black Hole Quencher 1 (BHQ1) or Cyanine 5 (CY5) and Black Hole Quencher 3 (BHQ3).

One fluorophore/quencher combination (FAM-BHQ-1) was included for the probe of each of the SARS-CoV-2 target genes. SARS-CoV-2 primer/probes were multiplexed in pair combinations with MS2 bacteriophage primers and probe (with a Cy5-BHQ3 fluorophore/quencher) to allow for detection of an internal extraction control. All primers and probes were HPLC purified and ordered from Metabion Ltd. (Germany).

A one-step RT-qPCR assay was used for all candidate target combinations, where each reaction well contained 6.25 µL TaqPath 1-Step RT-qPCR Master Mix, CG (ThermoFisher Scientific), 1 µL of 25X primer and probe mix, 7.75 µL molecular grade water and 10 µL total nucleic acid extract. Each RT-qPCR run contained the following controls: nucleic acid extracted SARS-CoV-2 positive control of the ancestral Wuhan strain (1:10,000 dilution; see Table 2 below); extracted L6 Buffer (negative control); molecular grade water (W4502-1L, Sigma-Aldrich; non-template control).

**Table 2.**
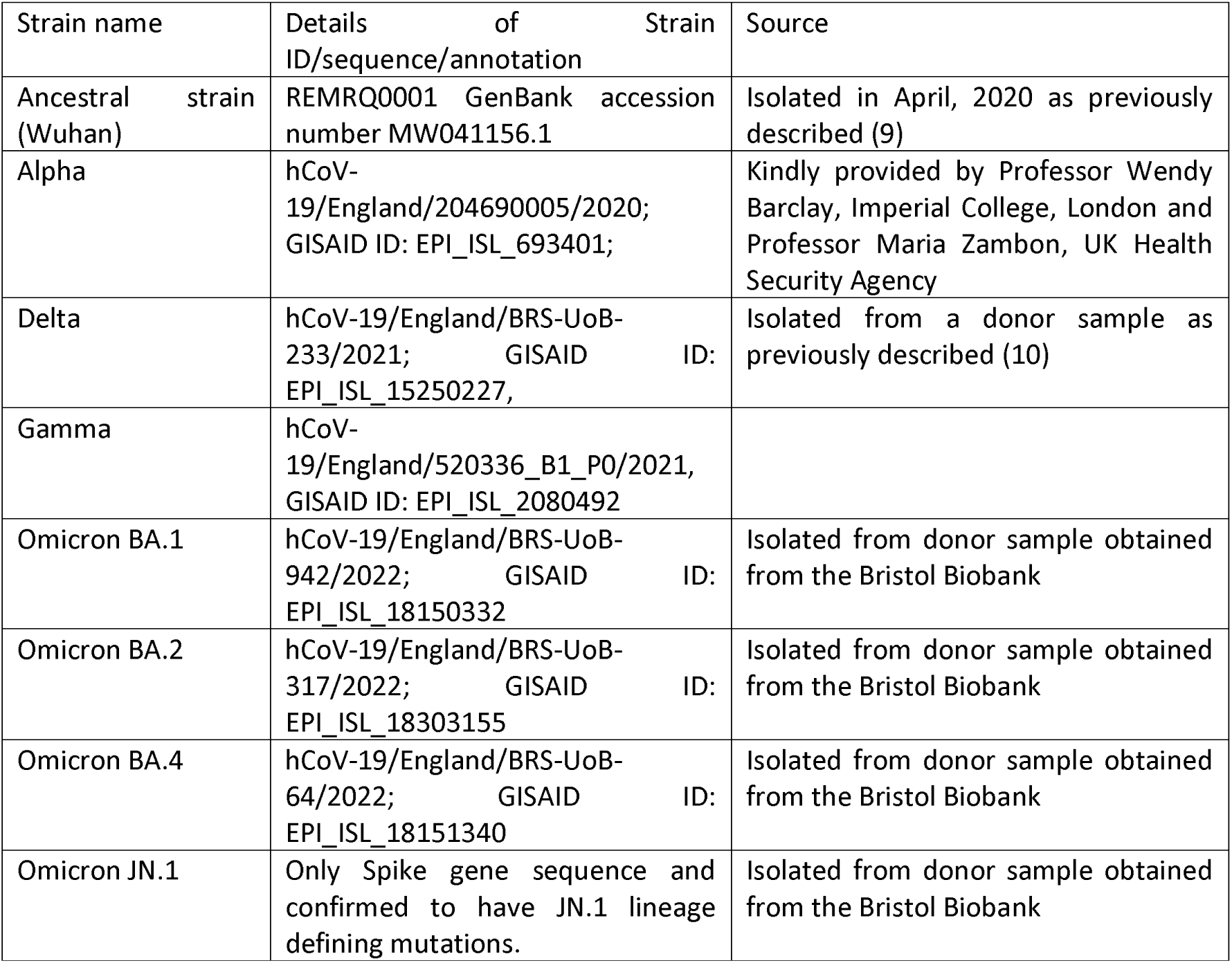
Details of sequences and sources of the various SARS-CoV-2 strains used to validate RT-qPCR assays and as positive control material.

Total nucleic acid and master mixes were pipetted using the automated QIAgility HEPA / UV Instrument (QIAGEN) before being run on the QuantStudio 7 Real-Time PCR System (Applied Biosystems). Thermal cycling consisted of 25°C for 2 mins, 50°C for 15 mins and 40 cycles of 95°C for 10 sec, 60°C for 30 sec (as per guidelines for TaqPath, ThermoFisher Scientific).

### Reaction Efficiencies and Limit of Detection

Reaction efficiencies were determined using dilutions of SARS-CoV-2 ancestral strain (Wuhan, see Table 1) RNA purified from tissue culture supernatant of SARS-CoV-2 infected cells as described previously (9). Limit of detection (LoD) was determined using RNA extracted from a commercial source of synthetic SARS-CoV-2 of known copy number (AccuPlex SARS-CoV-2 reference material, 4162 copies/ml, SeraCare Life Sciences Inc, USA). Purified RNA was diluted two-fold and between 8 and 16 replicates run at each dilution.

### Sensitivity and Specificity

Diagnostic sensitivity and specificity were determined using a panel of 60 SARS-CoV-2 positive and 59 SARS-CoV-2 negative samples kindly supplied by UKSHA Bristol and previously tested in their laboratories using a commercial kit (RealStar®SARS-CoV-2 RT-PCR Kit 1.0, Altona Diagnostics, Hamburg), which acted as the reference standard. To calculate confidence intervals for sensitivity and specificity values, the Clopper-Pearson method was used.

### SARS-CoV-2 Variants and Sequencing

To test the performance of the N6/E/MS2 and N2/N6/MS2 assays on different SARS-CoV-2 variants, viruses were obtained from multiple sources; details of these are provided in Table 2. In all cases, the specific strain ID was determined using Illumina mRNA sequencing (performed by staff at the Genomics Facility, University of Bristol) using the following kits/methods. A total volume of 8ul of total RNA sample was taken directly into the NEBNext® Artic SARS-CoV-2 Library Prep Kit Part Number E7650S/L (New England Biolands, Inc. Ipswich, MA) or the NEBNext® Artic FS SARS-CoV-2 Library Prep Kit Part Number E7658S/L followingthe manufacturer’s instructions without deviation.

Briefly, the protocol involved cDNA synthesis, followed by targeted cDNA amplification using two primer pair pools, A and B, to span the SARS-CoV-2 virus genome (primer schemes based on the original work of the ARTIC Network (https://github.com/joshquick/artic-ncov2019/blob/master/primer_schemes/nCoV-2019/V3/nCoV-2019.tsv). Primer schemes evolved throughout the running of this experiment and the latest version was used to represent current variants. Resulting amplicons from the two PCRs (A and B) per sample were pooled and cleaned up using a bead-based method (beads supplied in library preparation kit) before checking resulting amplicon size (1 in 10 dilution) on the Agilent TapeStation High Sensitivity DNA 1000 Screentape Assay (Agilent Technologies, Santa Clara, CA) on the Agilent 2200 TapeStation instrument. The amplicons were end-repaired prior to NEBNext ‘Adaptor for Illumina’ ligation, a further bead-based clean-up, PCR enrichment of the adaptor-ligated DNA to incorporate a unique dual-index barcode into each sample, and a final bead-based clean-up of the final libraries.

Final libraries were quantified using the Thermofisher High Sensitivity dsDNA Qubit assay (Thermofisher Scientific, Inc., Waltham, MA) and validated using the TapeStation (Agilent) High Sensitivity DNA1000 screentape assay on the Agilent 2200 TapeStation instrument. The libraries were normalised to 4nM, pooled equimolarly, and diluted to 10pM for sequencing on the Illumina MiSeq instrument using an Illumina Version 3, 600-cycle sequencing kit (Illumina Inc, San Diego, CA) and MiSeq Control Software Version 2.5.0.5. A spike-in of a PhiX control library Version 3 (Illumina Inc., San Diego, CA) at 10 percent was added to the sequencing run to assist with accurate base calling for a biased library. Primary data analysis was completed by on-board MiSeq Software (RTA Version 1.18.54.0) and FASTQ Creator (MiSeq Reporter Version 2.5.1.3). Data visualisation and QC metrics were assessed using Illumina BaseSpace Sequence Hub (basespace.illumina.com).

Positive samples identified from the field testing of BOTT OPS collected in 2020, were sequenced and strain-typed in Exeter by COG-UK using MinION sequencing as described in the published protocol (11).

### Data Analysis

Data analysis and presentation were performed with GraphPad Prism (v10). The limit of detection was defined as the copy number at which >95% of replicates were detected, as determined by fitting a 4 parameter logistic regression (least squares fit) on a plot of fraction samples positive against known copies of virus per RT-qPCR, and interpolating the value (and 95% confidence intervals where fraction positive = 0.95). RT-qPCR data from test samples were exported using either the QuantStudio Real-Time PCR Software v1.3 or the Qtower3. Any CT in the FAM channel for candidate assays indicated SARS-CoV-2 RNA had been detected. Cut-off values were determined using multiple repeats of serial dilutions of positive control material, and for all assays CT values <=35 were considered positive. Thresholds were set to 0.01 for SARS-CoV-2 N6 and E, which are indistinguishable since they both use the FAM channel and 0.01 for the MS2 bacteriophage used the Cy5 channel.

## Results

### Initial characterisation of N6/E/MS2 and N2/N6/MS2 RT-qPCR assays

We initially developed single plex assays to detect different genes from the SARS-CoV-2 genome, including two areas of the gene for nucleocapsid (N2, N6) and one each of envelope (E) and the RNA-dependent-RNA-polymerase (RdRp). Of these potential targets, primers binding to the E and N genes were found to provide the best efficiencies (data not shown). Two candidate dual target assays were developed, to improve on the sensitivity over single plex assays), including incorporation of control target (MS2) to assess for extraction, RT and qPCR performance. The candidate assays selected for further characterisation were: N6/E/MS2 and N2/N6/MS2.

We sought to determine limits of detection (LoD) and reaction efficiencies of the two candidate assays, using the ancestral (Wuhan) SARS-CoV-2. Both assays showed suitable efficiencies as multiplex assays and were compared to use of the targets in a single plex assay (Supplementary Figure 1). However, the N2/N6/MS2 assay had a greater reaction efficiency at 98% with R^2^= 0.995, compared to N6/E/MS2 at 95% (R^2^=0.998) (Supplementary Figure 1). The LoD was determined using multiple aliquots of samples with known copy numbers (Figure 1) and was defined as the copy number at which 95% of replicates were detected.

**Figure 1.**
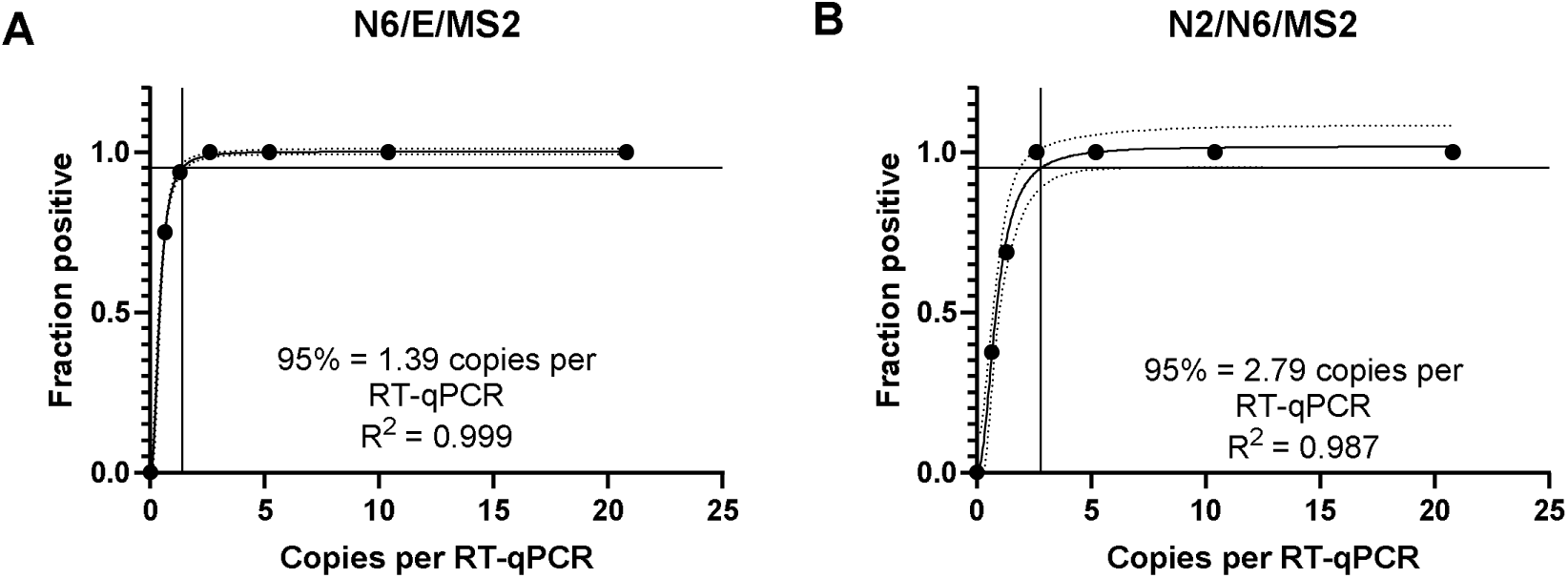
Limit of detection (LoD) was determined using RNA extracted from a commercial source of synthetic SARS-CoV-2 of known copy number (Accuplex SARS-CoV-2 reference material, 4162 copies/ml, Seracare Life Sciences Inc, USA). Purified RNA was diluted two-fold and between 8 and 16 replicates were run at each dilution. A) N6/E/MS2 and B) N2/N6/MS2 plots show the RNA copies per reaction on the axis and the % positive readings on each assay on the y axis with a curve plotted using 4 parameter logistic regression (least squares) with 90% confidence intervals also plotted. The LoD is defined as the copy number of virus at which 95% of replicates are detected as determined by interpolating the values when the fraction positive =0.95; these are stated in a panel within each plot, along with the R squared (R) for each assay.

The N6/E/MS2 assay displayed a lower LoD of 1.39 RNA copies per RT-qPCR (1.25-1.55, 90% CI) compared to N2/N6/MS2 at 2.79 (1.87 –6.35, 90% CI). At the highest dilution of virus at 0.65 copies per 10 µl reaction, the N6/E/MS2 assay detected almost double the number of positive results with very low viral copies compared to N2/N6/MS2 assay (75% compared to 37.5%).

### Sensitivity and Specificity

To determine assay performance of the two candidate assays, we performed a blind comparison with a gold standard dual target RT-qPCR kit which was commonly used at the time of assay development (2020), the Altona kit for monoplex detection of the S and E genes. Using a randomised set of 60 positive and 59 negative dual NPS and OPS samples provided by UKHSA Bristol (Severn Pathology, Southmead Hospital), we extracted RNA and tested the two candidate RT-qPCR assays. Both assays had high sensitivity for the positive samples (98.33% (59/60) for N6/E/MS2 and 96.7% (58/60) for N2/N6/MS2), and 100% (59/59) specificity, see Table 3).

**Table 3.** Diagnostic performance of candidate RT-qPCR assays. The sensitivity and specificity of the N6/E/MS2 and N2/N6/MS2 assays were compared to the results from the Altona assay (reference standard) used by the UKHSA.

| Candidate Assay | Positive Samples Detected as Positive out of Total | Negative Samples Detected as Positive out of Total | Sensitivity (95% CI) | Specificity (95% CI) |
| --- | --- | --- | --- | --- |
| N6/E/MS2 | 59/60 | 0/59 | 98.3% (91.06-99.96) | 100% (93.94-100) |
| N2/N6/MS2 | 58/60 | 0/59 | 96.7% (88.47-99.59) | 100% (93.94-100) |

Thus, both assays provided comparable diagnostic performance when compared to a widely used commercial assay. When comparing the two candidate assays, the lower limit of detection and slightly higher observed sensitivity led us to select the N6/E/MS2 assay as the better of the two in-house assays.

Use of N6/E/MS2 assay for detection of SARS-CoV-2 from samples collected in early 2020 The N6/E/MS2 PCR assay was used to detect SARS-CoV-2 in 1803 OPS samples with unknown SARS-CoV-2 infection status collected from healthy (i.e. asymptomatic) 16-18 year-old secondary school students in several sites across the UK between January and March 2020. These samples were collected as part of a large study of the impact of two meningococcal vaccines on throat colonisation in healthy teenagers (3). Of these samples, 4/1803 tested positive on the assay with each positive sample coming from a different site across the UK (Bristol, Cardiff, Glasgow and Maidstone) collected in March. The prevalence across the full study sampling period was 0.2% (95% CI 0.06 – 0.57) but in March the incidence was 0.45%; 2 of the 427 (0.47%, 95% CI 0.06% −1.7%) swabs taken during the week beginning 9th March were positive and 2 of the 459 (0.44%, 95% CI 0.05% −1.57%) swabs taken during the week beginning 16th March were positive (Supplementary Figure 2). To confirm detection of SARS-CoV-2 and identify strains, sequencing was performed on these samples by COG-UK in Exeter, which identified the samples as being of 3 lineages/clades of the ancestral Wuhan virus (12), results are shown in Table 4. These results suggest that the virus was in circulation at very low levels within secondary schools across England before the first lockdown period was implemented in March 2020.

**Table 4.**
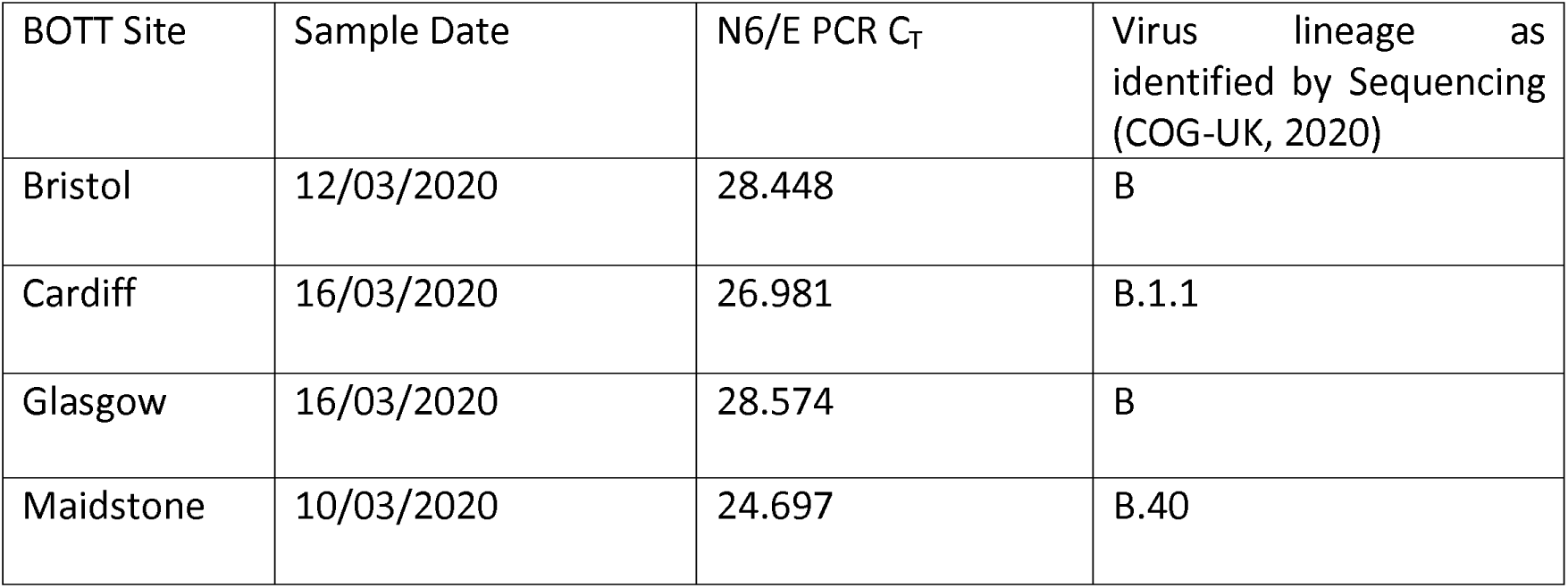
Details of the 4 OPS samples identified as positive from samples taken from children in March 2020 (total of n=1803 tested) as part of the Be on The Team (BOTT) study.

| BOTT Site | Sample Date | N6/E PCR C<sub>T</sub> | Virus lineage as identified by Sequencing (COG-UK, 2020) |
| --- | --- | --- | --- |
| Bristol | 12/03/2020 | 28.448 | B |
| Cardiff | 16/03/2020 | 26.981 | B.1.1 |
| Glasgow | 16/03/2020 | 28.574 | B |
| Maidstone | 10/03/2020 | 24.697 | B.40 |

### Detection of SARS-CoV-2 Variants

To confirm comparable detection of the key SARS-CoV-2 variants of interest (relating to different stages of the pandemic) in different respiratory samples, we initially performed 10-fold serial dilutions in culture media of four SARS-CoV-2 variants (Wuhan, Delta, Alpha and Omicron BA.1) which were important at the time. Samples were extracted using QIAsymphony and run on the N6/E/MS2 RT-qPCR assay, and were found to have good detection across the Ct range and suitable efficiency (Supplementary Figure 2). This was repeated more recently using Gamma and Omicron subvariants, including a virus of the JN.1 lineage; similar assay performance was observed for the variants (Figure 2B).

**Figure 2.**
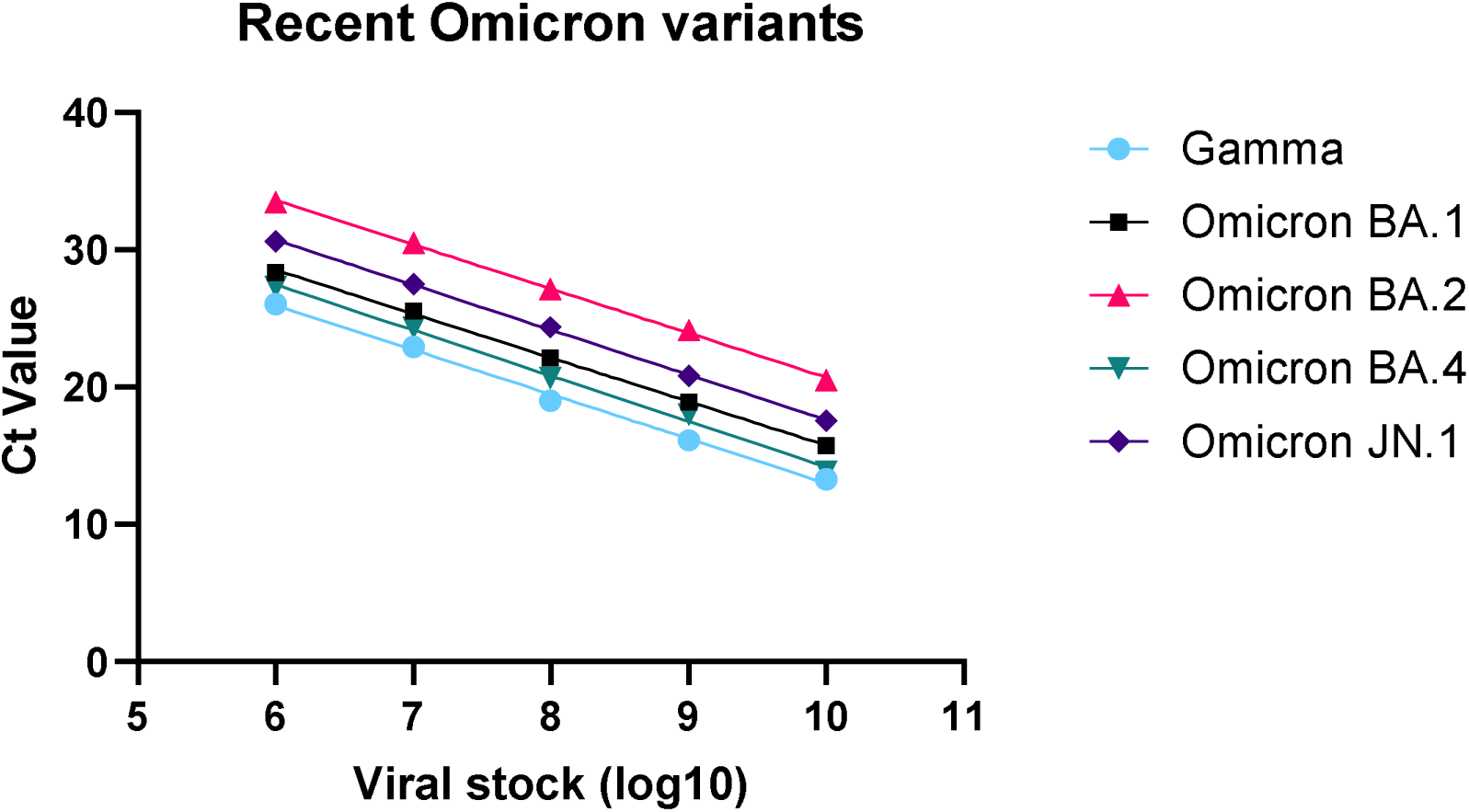
Detection of different SARS-CoV-2 Omicron and Gamma sub-variants with the N6/E/MS2 RT-qPCR assay. Detection of 10-fold serial dilutions of stock of SARS-CoV-2 Gamma strain (light blue dots), Omicron BA.1 (black squares), Omicron BA.2 (red triangles), Omicron BA.4 (green upside-down triangles), and Omicron JN.1 (dark blue diamonds) viruses. The X-axis represents the 10-fold dilution and the y axis the Ct value. The average Ct value for each dilution (3-6 replicates per dilution) is plotted along with the line of best fit after performing a simple linear regression.

The results demonstrate comparable performance for detection for each of the variants tested (also see supplementary Table 1), indicating applicability of the assay for a range of SARS-COV-2 variants representing several time periods since the start of the COVID-19 pandemic.

## Discussion

Quantitative RT-qPCR assays remain the gold standard approach for detection of SARS-CoV-2 in clinical samples. Here we present the development, performance characteristics and deployment of a novel dual-target SARS-CoV-2 RT-qPCR assay, N6/E/MS2. The assay demonstrates good accuracy for detection of SARS-CoV-2 in control and clinical samples as well as preserved ability to detect the major SARS-CoV-2 variants including the recently circulating and highly transmissible Omicron JN.1 subvariant (13).

Since the start of the COVID-19 pandemic, multiple commercial and in-house RT-qPCR assays have been developed for the detection of SARS-CoV-2. As others have reported, we found that sensitivity for detection can be improved by targeting multiple genes (14).. It is thus common for SARS-COV-2 RT-qPCR assays to be multiplex, with most currently-used commercial or recommended assays targeting a combination of the E, S, ORF1a and RdRp genes (14, 15). Different regions of the N gene are also common targets, such as in the assays published by the CDC (16). However, the N gene primers presented here are unique and offer high technical performance. With the use of M-Fold and primer3, we demonstrate how use of prediction software packages for the design of novel RT-qPCR assays can achieve high efficiencies and lead to good assay performance. We have also demonstrated the ability of the N6/E/MS2 assay to detect key SARS-CoV-2 variants of concern, including more recently circulating Omicron variant lineages. Conserved performance in these distinct variants of concern may relate to the choice of two distinct target gene regions (N6, E) which have remained relatively conserved in comparison with the S gene (17).

To field-test the assay in the early stages of the first UK wave of infections, ∼1800 samples from school age children collected in early 2020 (BOTT) were tested, and only four were found to be positive for SARS-CoV-2 (0.2%). Interestingly, these four positive samples were each identified from different sites, suggestive of minimal but widespread transmission of SARS-CoV-2 in UK in this age group at that time, despite the reported number of cases in the UK being only approximately 250 on March 10^th^ rising to ∼1,500 cases on March 15th i.e. when the samples were collected (18). This dispersion of isolated cases is supported by global transmission modelling studies that predict that community transmission was probably common in multiple parts of Europe as early as early January 2020 (19), and suggest that large numbers of asymptomatic infections went undetected at this time. Furthermore, sequencing data and modelling showed that there were multiple importation events of index cases into the UK in late February/Early March of 2020 from other parts of Europe (20). This is also compatible with our detection of isolated positive cases in 4 distinct locations in the UK more or less simultaneously. The sampling dates of the positive samples were close to the start of the introduction of a national lock down in the UK, on 23^rd^ March 2023 (18), which included school closures. However, given the low rate of infections detected in this cohort, it is unlikely that schools were driving a lot of the transmission at that time.

This study has some limitations. Whilst we have tested for sensitivity and specificity compared to a gold standard assay, these data were collected in the early stages of the pandemic, with the ancestral Wuhan virus, and we do not have assay comparison data for later variants. We included an internal RNA control (MS2) to monitor the efficiency of the extraction process. However, unlike other assays we did not include a human control probe to control for effective sample taking. Thus, some negative results observed may have been due to inadequate sampling.

In summary, the N6/E/MS2 assay is sensitive and specific for SARS-CoV-2 and has since been used to detect SARS-CoV-2 in respiratory samples collected as part of multiple studies, including a surveillance project in Bristol schools (“CoMMinS”) (21), and hospital and community-based surveillance studies of adult lower respiratory tract disease in Bristol, UK (“AvonCAP”) (22). As more recent SARS-CoV-2 variants have shown genetic drift, it is important that those using the assay perform regular checks on assay performance with current/recent variants.

## Data Availability

All data produced in the present study are available upon reasonable request to the authors

## Acknowledgements

We acknowledge both the Bristol Genomics Facility and COGG UK (Exeter) as the sequencing providers used for this research. We also acknowledge the Bristol Biobank for support in acquiring control samples for assay development.

