## Supplementary Information for "A sensitive multiplex RT-qPCR assay to detect SARS-CoV-2 in respiratory samples"

**Appendix 1**

BVC Microbiology and Molecular Labs:

Nadine King, Malak Eghleilib, Jessica Albuquerque Rosa, Mariella Ardeshir, Nellie Farhoudi, Chloe Farren, Ainhoa Rodriguez Pererira, William Healy, Caye Lisondra, Azizah Azis, Kajal Naukariya, Dylan Thomas, Louise Setter

Be on The Team Investigators:

Keith A Jolley, Karen Ford & Hannah Roberts (Oxford), Karen Palmer (Preston), Debbie Suggitt (Stockport), Nicola Pemberton (Wigan), Samantha Ray (Cardiff) Mandy Wootton (Cardiff), Shamez N. Ladhani (UKHSA), Daniel Owens & Katrina Cathie (Southampton), Simon Royal (The University of Nottingham Health Service), Neil Oldfield (School of Life Sciences, University of Nottingham), Roisin Ure, (Meningococcal Reference Lab, Glasgow), Jennifer Richards (Public Health Wales Microbiology), Rebecca Ramsay (Brighton), Samantha Thomson Hill (Bristol).

**Supplementary Figure 1**

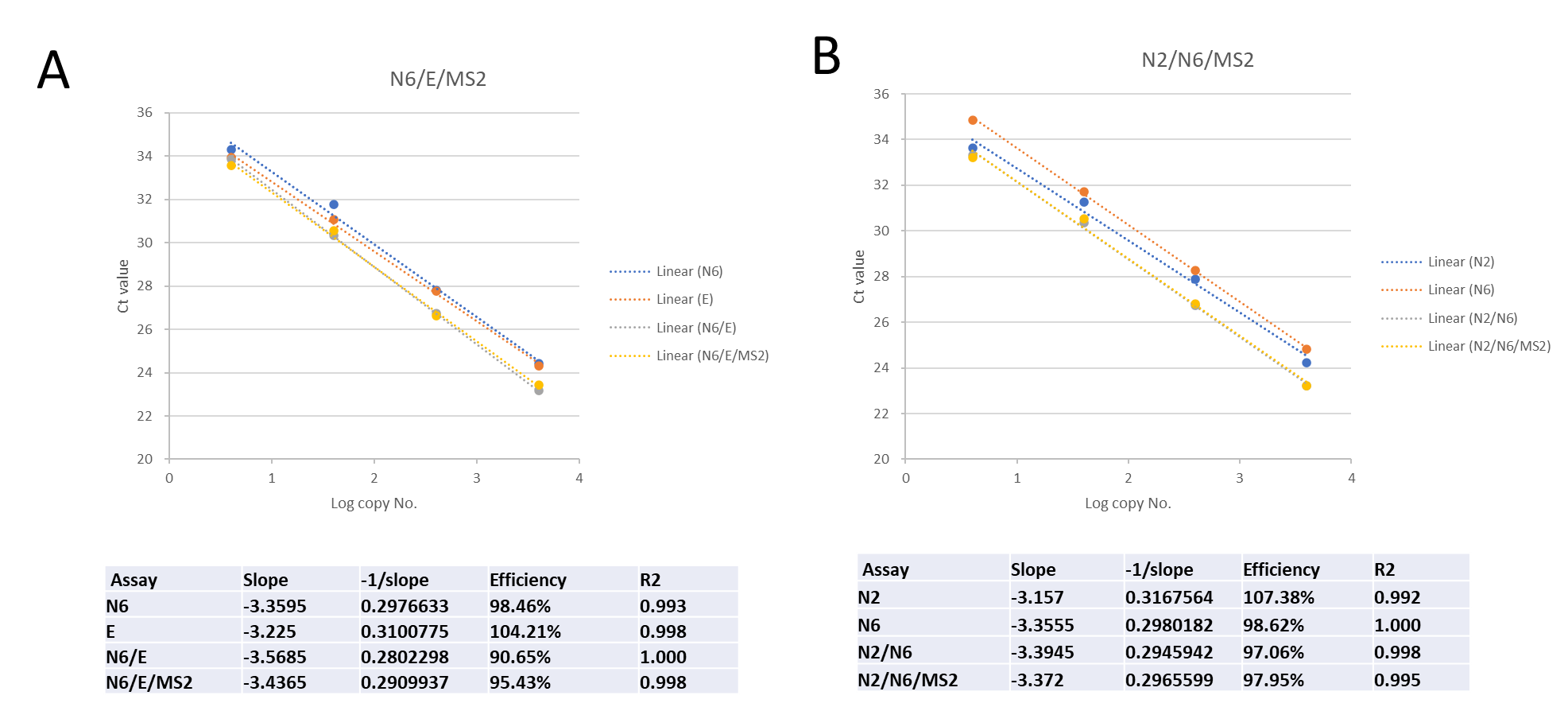

**Table S1**. RT-qPCR Assay efficiencies for detection of Ancestral strain SARS-CoV-2 using the monoplex N2, N6 or E assays, compared to the multiplex N6/E, N6/E/MS2 (panel A) or N2/N6 and N2/N6/MS2 assays (panel B). The plots show the average CT values for each assay and tables beneath report the slope, efficiency and R squared (R2) values for the assay, after simple linear regression.

**Supplementary Figure 2**

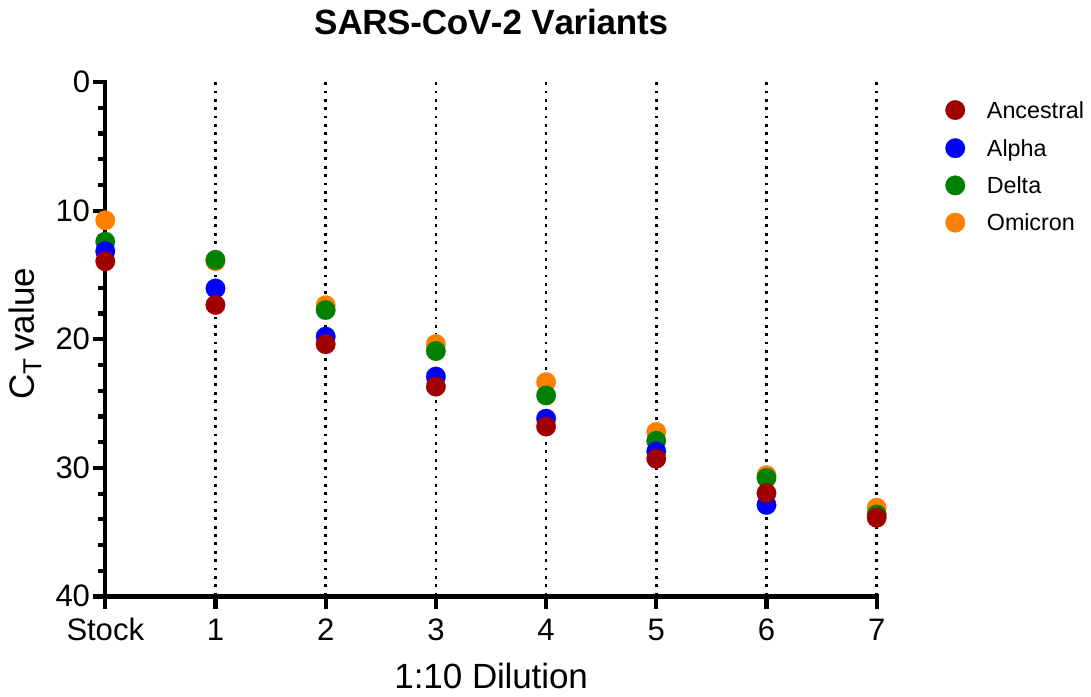

**Figure S2 –** Performance of N6/E/MS2 multiplex assay for detection of key SARS-CoV-2 variants of concern across a 10-fold dilution series using virus stock as that starting material. The extractions of each dilution and stock solutions were performed using the QIASymphony and replicates of each eluate were assayed using the optimised RT-qPCR protocol. Dots represent mean CT values from 3 replicates, for each 1:10 dilution.

**Supplementary Table 1**

**Table S1 –** Efficiency values for detection of different Gamma and Omicron sub-variants using the N6/E/MS2 RT-qPCR assays across 6 10-fold dilutions of virus stock. This table relates to the data presented in Figure 2.

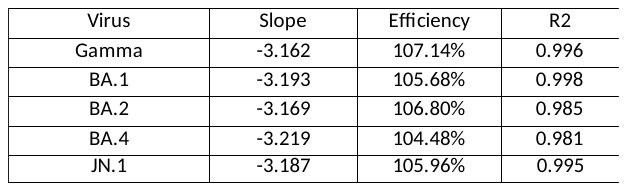
